# A machine learning-based approach to determine infection status in recipients of BBV152 whole virion inactivated SARS-CoV-2 vaccine for serological surveys

**DOI:** 10.1101/2021.12.16.21267889

**Authors:** Prateek Singh, Rajat Ujjainiya, Satyartha Prakash, Salwa Naushin, Viren Sardana, Nitin Bhatheja, Ajay Pratap Singh, Joydeb Barman, Kartik Kumar, Raju Khan, Karthik Bharadwaj Tallapaka, Mahesh Anumalla, Amit Lahiri, Susanta Kar, Vivek Bhosale, Mrigank Srivastava, Madhav Nilakanth Mugale, C.P Pandey, Shaziya Khan, Shivani Katiyar, Desh Raj, Sharmeen Ishteyaque, Sonu Khanka, Ankita Rani, Promila, Jyotsna Sharma, Anuradha Seth, Mukul Dutta, Nishant Saurabh, Murugan Veerapandian, Ganesh Venkatachalam, Deepak Bansal, Dinesh Gupta, Prakash M Halami, Muthukumar Serva Peddha, Gopinath M Sundaram, Ravindra P Veeranna, Anirban Pal, Ranvijay Kumar Singh, Suresh Kumar Anandasadagopan, Parimala Karuppanan, Syed Nasar Rahman, Gopika Selvakumar, Subramanian Venkatesan, MalayKumar Karmakar, Harish Kumar Sardana, Animika Kothari, DevendraSingh Parihar, Anupma Thakur, Anas Saifi, Naman Gupta, Yogita Singh, Ritu Reddu, Rizul Gautam, Anuj Mishra, Avinash Mishra, Iranna Gogeri, Geethavani Rayasam, Yogendra Padwad, Vikram Patial, Vipin Hallan, Damanpreet Singh, Narendra Tirpude, Partha Chakrabarti, Sujay Krishna Maity, Dipyaman Ganguly, Ramakrishna Sistla, Narender Kumar Balthu, A Kiran Kumar, Siva Ranjith, B Vijay Kumar, Piyush Singh Jamwal, Anshu Wali, Sajad Ahmed, Rekha Chouhan, Sumit G Gandhi, Nancy Sharma, Garima Rai, Faisal Irshad, Vijay Lakshmi Jamwal, MasroorAhmad Paddar, Sameer Ullah Khan, Fayaz Malik, Debashish Ghosh, Ghanshyam Thakkar, S K Barik, Prabhanshu Tripathi, Yatendra Kumar Satija, Sneha Mohanty, Md. Tauseef Khan, Umakanta Subudhi, Pradip Sen, Rashmi Kumar, Anshu Bhardwaj, Pawan Gupta, Deepak Sharma, Amit Tuli, Saumya Ray chaudhuri, Srinivasan Krishnamurthi, L Prakash, Ch V Rao, B N Singh, Arvindkumar Chaurasiya, Meera Chaurasiyar, Mayuri Bhadange, Bhagyashree Likhitkar, Sharada Mohite, Yogita Patil, Mahesh Kulkarni, Rakesh Joshi, Vaibhav Pandya, Sachin Mahajan, Amita Patil, Rachel Samson, Tejas Vare, Mahesh Dharne, Ashok Giri, Sachin Mahajan, Shilpa Paranjape, G. Narahari Sastry, Jatin Kalita, Tridip Phukan, Prasenjit Manna, Wahengbam Romi, Pankaj Bharali, Dibyajyoti Ozah, Ravi Kumar Sahu, Prachurjya Dutta, Moirangthem Goutam Singh, Gayatri Gogoi, Yasmin BegamTapadar, Elapavalooru VSSK Babu, Rajeev K Sukumaran, Aishwarya R Nair, Anoop Puthiyamadam, PrajeeshKooloth Valappil, Adrash Velayudhan Pillai Prasannakumari, Kalpana Chodankar, Samir Damare, Ved Varun Agrawal, Kumardeep Chaudhary, Anurag Agrawal, Shantanu Sengupta, Debasis Dash

## Abstract

Data science has been an invaluable part of the COVID-19 pandemic response with multiple applications, ranging from tracking viral evolution to understanding the effectiveness of interventions. Asymptomatic breakthrough infections have been a major problem during the ongoing surge of Delta variant globally. Serological discrimination of vaccine response from infection has so far been limited to Spike protein vaccines used in the higher-income regions. Here, we show for the first time how statistical and machine learning (ML) approaches can discriminate SARS-CoV-2 infection from immune response to an inactivated whole virion vaccine (BBV152, Covaxin, India), thereby permitting real-world vaccine effectiveness assessments from cohort-based serosurveys in Asia and Africa where such vaccines are commonly used. Briefly, we accessed serial data on Anti-S and Anti-NC antibody concentration values, along with age, sex, number of doses, and number of days since the last vaccine dose for 1823 Covaxin recipients. An ensemble ML model, incorporating a consensus clustering approach alongside the support vector machine (SVM) model, was built on 1063 samples where reliable qualifying data existed, and then applied to the entire dataset. Of 1448 self-reported negative subjects, 724 were classified as infected. Since the vaccine contains wild-type virus and the antibodies induced will neutralize wild type much better than Delta variant, we determined the relative ability of a random subset of such samples to neutralize Delta versus wild type strain. In 100 of 156 samples, where ML prediction differed from self-reported uninfected status, Delta variant, was neutralized more effectively than the wild type, which cannot happen without infection. The fraction rose to 71.8% (28 of 39) in subjects predicted to be infected during the surge, which is concordant with the percentage of sequences classified as Delta (75.6%-80.2%) over the same period.

## Introduction

Mathematical and statistical methods have not only proven helpful to model epidemiological data but also handled the ever-growing host-pathogen data to combat COVID-19 effectively. So far diverse COVID-19 disease outcome models have been developed using electronic health records, epidemiological and symptoms data (Estiri et al., 2021; Gupta et al., 2020; Zoabi et al., 2021). Transmission rate and viral load kinetics have been studied using mathematical models on vaccination data (Singanayagam et al., 2021). Antibody kinetic analysis found 36% anti-S antibodies after one year of infection in a serological setting (Pelleau et al., 2021).

Modeling based on RT-PCR based inputs relies on infection status at that time but misses the previous infection history for which serosurvey-based data is instrumental. Latter also comes with some challenges e.g. waning immunity with time. Nevertheless, serological studies provide complementary information about the infection history of the individual, especially when reports surface mentioning that previous infection is helpful to generate the anti-SARS-CoV-2 antibodies apart from vaccination (Kang et al., 2021). Considering these findings, it is important to ascertain the previous infection, especially in serological analyses. Moreover, serological data can identify asymptomatic infection, which is not possible using conventional methods, namely RT-PCR.

Inactivated whole virion vaccines have been showing encouraging results in protection against the COVID-19 (Ella et al., 2021) and has been approved by WHO on November 3^rd^, 2021, under the Emergency Use Listing (EUL) category. Recent pilot studies have shown Covaxin efficacy based on neutralization and antibody response against variants of SARS-CoV-2 and it was reported that Covaxin is effective against new emerging variants (Sapkal et al., 2021a; Sapkal et al., 2021b; Yadav et al., 2021a; Yadav et al., 2021b). Recently, Covaxin was observed to have a protection efficacy of 47% in previously uninfected individuals, after two doses for symptomatic presentation (Desai et al., 2021).

While the vaccine efficacy is determined by the RT-PCR test in general, the same test is limited to symptomatic cases, and contact tracing. However, asymptomatic infections are largely ignored. In contrast, serology-based assessment of vaccine efficacy becomes more pertinent in the context of the general population especially where RT-PCR testing is infrequent and viral load is insignificant (Hodgson et al., 2021; Lee et al., 2020). Vaccines targeted specifically to the spike protein do not pose a problem in detecting positive infection since positive Anti-NC is taken as a marker of infection. However, for the whole virion vaccine, Anti-NC is induced by the vaccine itself which creates challenges to identify infection status and hence ascertaining the vaccine protection efficacy.

To address this issue, we developed a hybrid machine learning approach based on serological values to Anti-NC and Anti-S along with other demographics and infection history in Covaxin recipients to determine infection status and thus protection efficacy of the vaccine. To develop and validate our method, we used the serosurvey data from CSIR Cohort, a longitudinal cohort that was developed to assess the disease burden across India and to assess the stability of antibodies during post-infection/vaccination (Dhar et al., 2021; Naushin et al., 2021). Population-based cohorts could help to determine the confident estimates of vaccine efficacy with a heterogenous accommodation of larger geographical regions. To robustly ascertain the infection status, we took the consensus of two approaches namely an unsupervised clustering approach and a supervised SVM-based approach followed by an ensemble model for final infection prediction. We also validated the outcomes of ML models using the surrogate Virus Neutralization Test (sVNT). To the best of our knowledge, this is the first work to predict the infection status and protection efficacy of Covaxin-vaccinated individuals based on serological analysis and clinical history.

## 2 Methods

### 2.1 Data/cohort description

The samples analyzed in this study were from a longitudinal cohort of staff, students, and their family members belonging to 43 CSIR laboratories and centers of the Council of Scientific and Industrial Research (CSIR) spread across India (CSIR Cohort; (Naushin et al., 2021)) who had taken one or two doses of Covaxin. The longitudinal cohort study was approved by the Institutional Human Ethics Committee of CSIR-IGIB vide approval CSIR-IGIB/IHEC/2019-20. To date, samples have been collected from this cohort in three phases: between June-September 2020 (Phase 1; P1), between January 2021-March 2021(Phase 2; P2), and May-August 2021 (Phase 3; P3). Phase 3 was incidentally bracketed with the COVID-19 second wave (April 2021-August 2021) in the country and dominated by the Delta variant of SARS-CoV-2 (Dhar et al., 2021).

All subjects participated voluntarily and filled an online questionnaire form which included information on the date of birth, gender, blood group, type of occupation, comorbidities such as Diabetes, Hypertension, Cardiovascular Diseases, etc, diet preferences, mode of travel, symptomatology, vaccine status, hospitalization if any, post-vaccine symptomatology if any. These forms were then downloaded in MS-Excel data format and merged with registration forms filled at the time of sample collection based on unique IDs. Blood samples (6 mL) were collected for each subject in an EDTA-coated vacutainer and centrifuged at 1800 g for 15 minutes at 4°C. Separated plasma was stored at -80°C until used to assess antibodies against recombinant protein representing Nucleocapsid (NC) and Spike (S) antigens of SARS-CoV-2 using Elecsys Anti-SARS-CoV-2 kits (Roche Diagnostics) based on Electro-chemiluminescence Immunoassay (ECLIA) according to manufacturer’s procedure. Individuals with a Cut-off index (COI) value of > 1.0 and a value of > 0.8 U/mL were considered to be positive for Anti-NC and Anti-S antibodies, respectively. Wherever necessary, samples were appropriately diluted for the Anti-S antibody measurements (Dhar et al., 2021).

### 2.2 Algorithm Development

For a supervised and an unsupervised approach; Phase 3 (P3) samples with sero history information in Phase 1 and/or Phase 2 (P1/P2), were selected. In P3, people had self-reported if they had been infected with the SARS-CoV-2 virus as confirmed through RT-PCR or had symptoms, but no RT-PCR was done (“not confirmed”). These subjects were categorized as “self-reported infected”. Similarly, people without any symptoms and did not feel that they were infected (some of them also reported RT-PCR negative) were categorized as ‘self-reported not infected’. Those who were seropositive in P1/P2 were considered infected henceforth. This was an important consideration as P2 negative samples would have helped us ascertain protection against Delta for that was the period when Delta was the predominant variant. Finally, 1063 out of 1823 individuals qualified as input data for downstream analyses (414 infected and 649 not infected samples). The flow diagram for the algorithm to final prediction pipeline is depicted in **Figure 1**.

**Figure 1:**
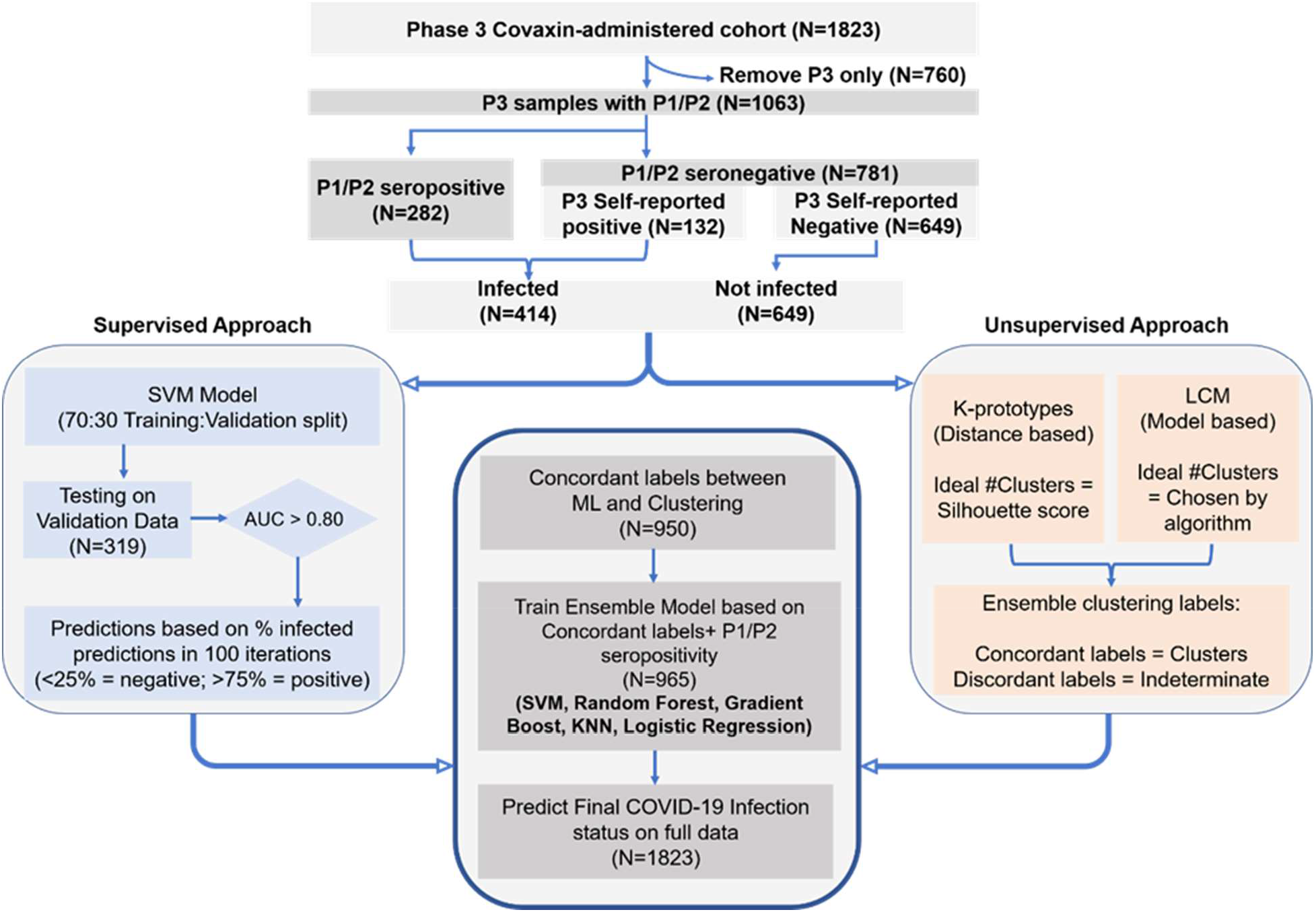
Overall workflow of the study used to identify COVID-19 infection status. Using a consensus of supervised (machine learning) and unsupervised (clustering) approaches, COVID-19 Infection status was ascertained in 1063 individuals who provided samples in Phase 3 (P3) and also in Phase 1 or Phase 2 (P1/P2). The final ensemble model was used to predict the COVID-19 infection status for all Covaxin administered individuals in P3.

The elements of input data included serological assays - qualitative Anti-NC (COI) and quantitative Anti-S (U/mL) values of samples along with age, gender, number of vaccination doses, and days since the last vaccination. Further, COI and U/mL values were log10 transformed and distribution was calculated using density plots. Days since the last vaccination was also log10 transformed for supervised ML algorithms. The supervised approach included SVM-based model development over 100 iterations with random splitting of 70:30 ratio of training and testing datasets where final labels were assigned based on the consensus of 100 iterations. Unsupervised clustering itself consisted of two orthogonal approaches, concordance of which is carried forward as cluster 1 and cluster 2. The consensus of the supervised and the unsupervised approaches in addition to P1/P2 seropositive individuals is further used for the final ensemble ML model to predict the full dataset (**Figure 1**). Both the approaches are explained in the following sections.

#### 2.2.1 Unsupervised clustering approach

For robustness of the unsupervised analysis, we used two separate approaches namely K-prototype and LCM-based approaches. K-prototype (Szepannek, 2018) is a distance-based mixed-mode (Numerical and Categorical) clustering method that uses distance as a measure to define the clusters while VarSelLCM (Marbac and Sedki, 2019) is a mixed-mode model-based method that uses probabilities derived from a model defined using Latent Class Analysis for defining the clusters. The number of clusters best describing the data was first established based on silhouette score (for K-prototype algorithm), while the VarSelLCM clustering algorithm chooses the best number of clusters automatically. The concordance of cluster assignment between the two methods was ascertained as Cluster 1 and Cluster 2 and discordant labels were labeled ‘Undetermined’. Cluster 1 was predominantly the region which included self-reported infected samples while cluster 2 had a wider dispersion and included self-reported non infected samples.

#### 2.2.2 Supervised machine learning

Input data features were preprocessed for feature scaling using the standard scaler library in scikit-learn (Pedregosa et al., 2011), and data was split in a 70:30 ratio for training and testing sets, respectively. Briefly, to ensure that each data point gets a chance to be treated as test data multiple times, 70% (training set) of 1063 entries were trained on the machine learning model and the rest 30% (319) was used as (blind set) for validation. In every iteration, data splitting from 1063 samples was randomized i.e. no two iterations had identical data as training set or blind set for testing. SVM algorithm (uses hyperplane to separate two classes) was used for model development on 70% of training data with 5-fold cross-validation technique where grid search was used for best parameters selection and validated on 30% blind dataset. 100 such iterations each with an area under the receiver operating characteristic curve (AUROC) of greater than 0.80 was performed. The robustness of ML prediction was ensured by iterating the whole pipeline 100 times. A sample was considered infected or not infected only if they were classified as such in at least 75% of the total times predicted, else they were marked as indeterminate.

#### 2.2.3 Ensemble Clustering

The outputs of the unsupervised and the ML-based analysis were then compared. We found the concordance between ML-based infected samples with Cluster 1 of consensus clustering approach (labeled infected class) and ML-based uninfected samples with Cluster 2 of consensus clustering approach (labeled uninfected class). About 1.8% (19/1063) who had a previous positive history of seropositivity but classified as Not infected/Cluster 2 or Indeterminate by both the methods were reassigned to be positive. These consensus samples were then used to develop a 5 ML-classifiers (SVM, logistic regression, K-Nearest Neighbors, Random Forest, and Gradient Boost) based ensemble model. This model was then used to predict the infection status of all the samples.

### 2.3 Validation of potential asymptomatic samples with neutralizing antibodies

The final model was validated by testing a few samples for their neutralizing activity against Delta infection. For this, a surrogate neutralization was performed against both wild type and Delta Receptor-Binding Domain (RBD) using GenScript cPass SARS-CoV-2 surrogate Neutralization Antibody detection kit -sVNT assay (GenScript, USA) as per manufacturer’s instruction (Naushin et al., 2021). Since the vaccine was developed against the wild-type variant, it is expected that the neutralization will be higher if the neutralizing activity is due to vaccination or infection with wild-type virus or both.

However, if a person has been infected with Delta variant then the neutralizing activity against the Delta RBD will be higher or equal to that of wild-type RBD. Where required, samples were diluted five times. A neutralization of 30% or more in undiluted samples was considered to be positive. To determine the Delta Infection status of the remaining, we first determined the standard error between technical replicates of the samples at various inhibition percentages (%age). Using this we decided on the Lower bound (LB) of the Wild-type inhibitions% and Upper bound (UB) of the Delta-type inhibition %age as Inhibition %age minus, and plus twice the average standard error respectively. Using these criteria, we called any sample with UB of Delta > LB of Wild-type to be a possible Delta Infection.

### 2.4 Vaccine efficacy calculation

The unvaccinated group comprised 910 participants who were negative in Phase 2 of the CSIR cohort study. Of these 561 had become seropositive on sample collection in Phase 3 of sample collection, while 349 remained uninfected. Of 187 subjects who took Covaxin from January-May 2021 and were negative in Phase 2; 52 were predicted to be positive by the ML algorithm. Thus, protection was calculated using the following formula;

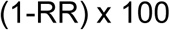

where RR is the relative risk for the vaccinated group to the unvaccinated group.

### 2.5 Statistical Analysis

We used the R software package version 3.5.1 for the data curation, management, and clustering analysis. We used the Python library, namely, Scikit-learn (version 0.24.1) for predictive modeling.

## 3 Results

### 3.1 Data/cohort details

Blood samples from 1823 Covaxin administered participants belonging to phase 3 (22 May 2021-09 Aug 2021) of CSIR-cohort were processed for the Anti-NC and Anti-S antibody assays. Location-wise distribution of individuals has been described in **Supplementary Table S1**. The details of baseline characteristics are presented in **Supplementary Table S2**. Out of the 1823 individuals, 772 had taken one dose while 1051 had taken two doses of Covaxin. Of these; 789 and 792 individuals have provided samples in P1 and P2, respectively (**Figure 2A)**.

**Figure 2:**
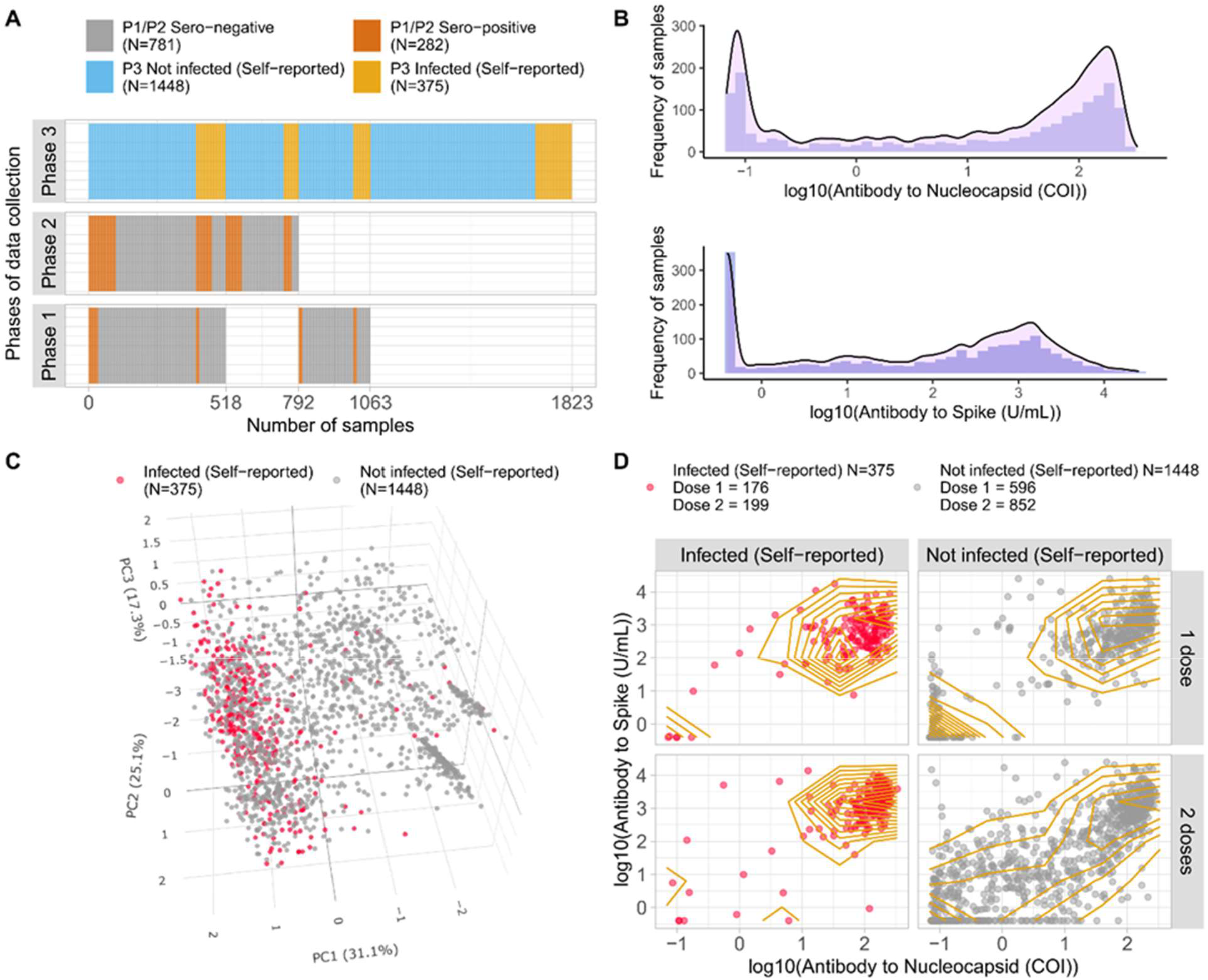
A): Sample distribution and overlap among three phases [P1 (June-November, 2020), P2 (December, 2020-April, 2021), P3 (May-August, 2021)] of CSIR Cohort of Covaxin administered individuals (N=1823), B): Distribution of Antibodies to Nucleocapsid (COI) and Spike (U/mL) in the form of density histograms of 1823 individuals, C): PCA plot of 1823 Covaxin administered individuals based on six features including COI, U/mL, age, gender, days since last vaccination, and the number of doses. COVID-19 self-reported infection is depicted in red color, D): Sample distribution stratified via self-reported COVID-19 infection status and doses taken (N=1823). Density-based contours indicate the presence of two subgroups amongst both in 1 dose and 2 doses administered self-reported not infected individuals.

The preliminary density distribution in terms of Anti-NC and Anti-S antibodies shows bimodal distribution indicating the presence of two subgroups (**Figure 2B**). Further, PCA transformation of the full data, including similar parameters given for supervised/unsupervised approaches (see methods), was analyzed using the top three principal components, PC1, PC2, and PC3, which explained 31.1%, 25.1%, and 17.3% of the variance of the data respectively (**Figure 2C, Supplementary Figure S1**). The PCA plot shows the distribution of self-reported infected (red color) and uninfected individuals (grey color). The self-reported infected individuals formed a cluster, while the self-reported negative individuals were spread throughout. It should be noted that although confirmed positive RT-PCR or presence of symptoms most likely indicate infection, a negative PCR test or absence of symptoms does not rule out infection since the RT-PCR test is positive only during a short window of infection (Mallett et al., 2020). Also, we and others have earlier shown that a large proportion of individuals infected with SARS-CoV-2 are asymptomatic (Bai et al., 2020; Naushin et al., 2021; Nishiura et al., 2020; Oran and Topol, 2020). This was also observed when the Anti-NC and Anti-S antibody levels were plotted for self-reported positive and negative subjects (**Supplementary Figure S2**). This was irrespective of the vaccine doses since self-reported positive subjects with one (N=176) and two doses (N=199) had similar levels of antibodies forming a single cluster, while those of self-reported negative with one (N=596) or two doses (N=852) showed two distinct clusters and had large contours of distribution (**Figure 2D**).

The observations are suggestive of a probable undiagnosed infection among the self-reported uninfected individuals. To ascertain the extent of infection in these individuals we developed a two-step workflow consisting of unsupervised clustering and supervised SVM-based ML model in parallel followed by an ensemble model whose inputs consisted of consensus classifications obtained from the two methods in step 1 (**Figure 1**).

#### 3.2.1 Unsupervised clustering identifies two clusters

1063 participants whose prior serology status at phase 2 was available were subject to K-prototype and VarSelLCM clustering algorithms. To consider all relevant confounders while creating the clusters, log10 of Antibody to Nucleocapsid, log10 of Antibody to Spike, age, gender, number of doses, and days since last vaccination for each person were provided as input to the algorithms. Statistical methods in respective clustering methods identified the ideal number of clusters to be two (see methods). Between the two methods, there was a concordance of 96.05% (1021/1063) as shown in the consensus clustering output while 42 (3.95%) discordant samples were mainly concentrated at the junction of the two clusters (black dots, **Figure 3A**). Of the 236 self-reported infected samples 93.2% (220/236) were found to be in cluster 1, while of the 827 self-reported uninfected 50.4% (417/827) were in cluster 2.

**Figure 3:**
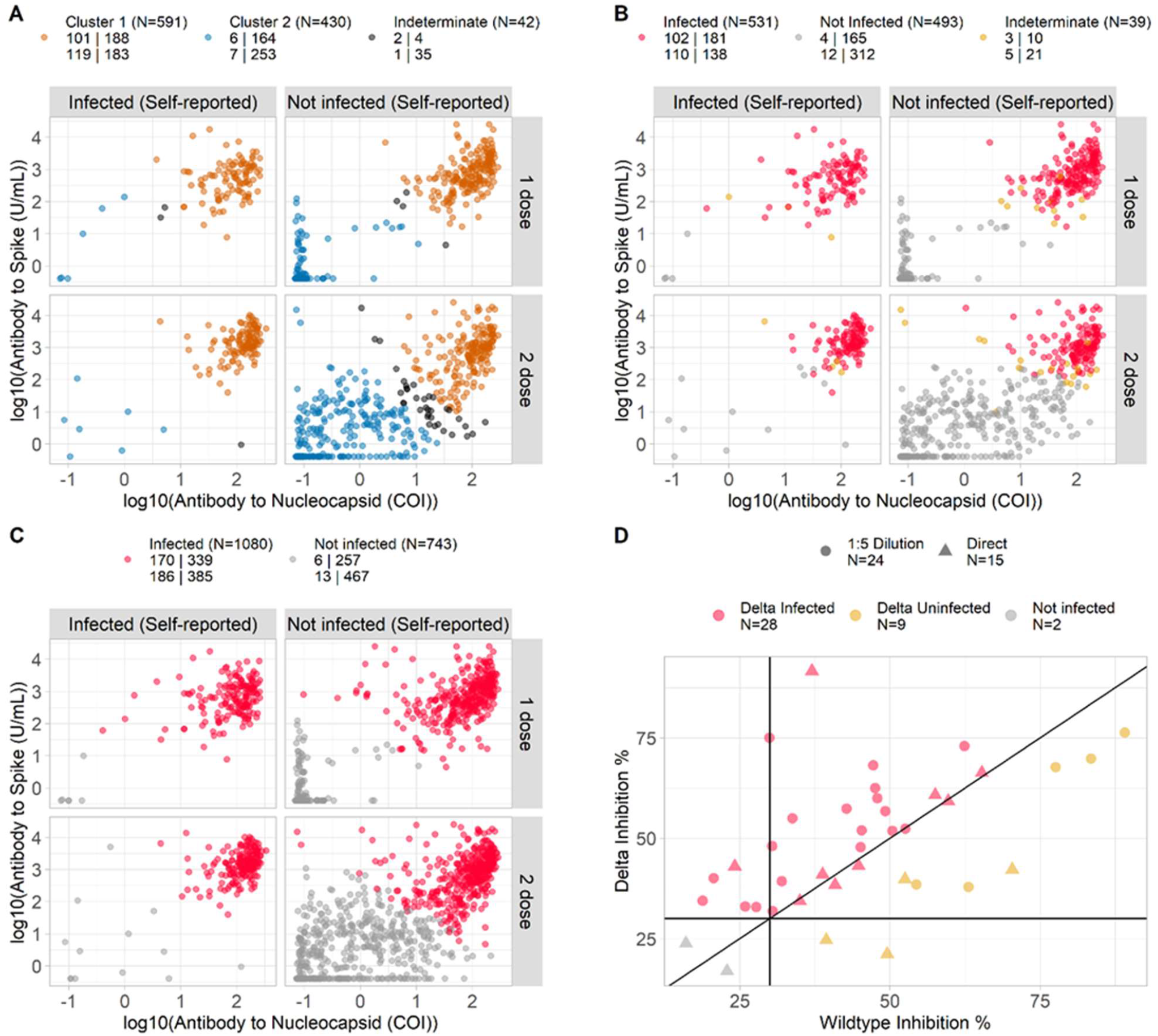
A): Consensus clustering with k-prototype and LCM methods (N=1063). Light Brown and blue colour represent concordance between two clustering approaches for Cluster 1 and Cluster 2 respectively. The black color represents discordance between the two methods, hence indeterminate; B): Supervised machine learning (SVM method) based prediction of the infection status (N=1063), further stratified via self-reported COVID-19 infection status and the number of vaccine doses; C): Ensemble ML model-based prediction of COVID-19 infection in all individuals (N=1823), further stratified via self-reported infection status and the number of vaccine doses; D): Phase 2 seronegative subjects who gave samples in Phase 3 analyzed using a surrogate virus neutralization assay (sVNT) and predicted to be Infected by Ensemble model (N=39). 71.8% of samples predicted to be Infected by Ensemble were found to be Delta Infected utilizing a variant-specific sVNT assay. Delta Infected was labelled when Delta Inhibition % > WT Inhibition % with a margin based on standard error. Delta Not infected were labelled when samples processed without dilution had less than 30% inhibition. All other data points were labeled Delta Uninfected.

### 3.2.2 Supervised ML to predict infection rate

In parallel to the clustering approach, we also developed an SVM-based ML model. For this, as before, we used the 1063 samples of which 414 samples which either had an earlier (P1/P2) seropositive history or self-reported infection status in P3 were considered to be infected and 649 samples with previous seronegative history and self-reported uninfected status at P3 were considered to be uninfected. SVM-based ML model with 5-fold cross-validation was built on 70% of the randomly selected data (N=744) while the same model was tested on 30% of the data (N=319) as mentioned in the methods. If the prediction on blind data was >0.80 AUROC, only then that prediction iteration was considered valid, and this step was repeated for 100 valid iterations. This process reinforced the robustness of the pipeline where each sample was tested at least 16 times (ranging from 16 - 44).

The average accuracy and AUROC obtained after 100 iterations were 81.3% and 0.88, respectively (**Supplementary Figure S3**). The predicted labels over 100 iterations were plotted as sigmoid curves, and samples that were predicted to be positive (531/1063; 49.95%) or negative (493/1063; 46.37%) in at least 75% of the models were labeled infected and uninfected, respectively, while the rest (39/1063; 3.66%) were indeterminate (**Figure 3B**).

#### 3.2.3 Ensemble ML model results

We calculated the concordance of ML and clustering approaches with their respective final 1063 labels. Earlier, cluster 1 and cluster 2 were enriched in infected and not infected individuals respectively; we found 526 individuals predicted to be Infected by ML and were present in Cluster 1, while 424 individuals were predicted to be uninfected by ML and present in Cluster 2, overall leading to 89.3% concordance. There was 1 sample indeterminate in both cases. As expected, P1/P2 seropositive samples (N=282) were mostly enriched in the concordant infected panel. The 19 P1/P2 seropositive samples (6.7%) that were assigned negative or indeterminate in the consensus model were reassigned positive (infected) to build the final model as they were infected for sure at one point in time. Out of these 19 samples, 4 were already classified as Concordant Not Infected but were reclassified as Infected based on their P1/P2 seropositivity status. **Supplementary Figure S4** shows one to one comparison of three categories of both ML and clustering approaches.

For the final ML ensemble model, we picked concordant (infected, red-colored (N=545) as well as not infected, grey-colored (N=420)) samples. An ensemble model based on 965 concordant samples was developed using 5 ML algorithms namely SVM, RF, Gradient Boost, KNN, and Logistic Regression. Using this final model, we assigned infection status for all the 1823 subjects who had taken Covaxin (**Figure 3C**). The ensemble model was able to correctly recognize as Infected 95% (356/375) and 99% (279/282) of the Self-reported infected and P1/P2 seropositive, respectively. Further, the model also assigned 50% (724/1448) of the self-reported negative samples as infected. The prediction on blind 858 individuals stratified by self-reported infection status and the number of doses is represented in **Supplementary Figure S5**.

### 3.3 Validation

The final predictions of the ensemble model were validated using surrogate virus neutralization (sVNT) assays as described in the methods section. All the vaccines have been developed against the wild-type SARS-CoV-2 strain, and hence it is expected that neutralization against wild-type RBD will be higher if it is due to vaccination or prior infection with wild-type variant. However, if a person has been infected with the Delta variant, which was predominant (75-80% of all infections) during this period across the country, then the neutralization against Delta RBD will be equal to or greater than wild type RBD. We found that 64.1% (100/156) individuals who self-reported uninfected but were predicted to be infected by the ensemble model, neutralized Delta RBD greater than or equal to that of wild type, suggesting that these individuals were probably infected by Delta variant although they were asymptomatic (**Supplementary Figure S6**). Of these, 71.8% (28/39) who were seronegative in P2 i.e., before the second wave and self-reported negative, were found to have higher Delta neutralization (**Figure 3D**), which was similar to the frequency of Delta infection across the country.

### 3.4 Vaccine efficacy results

Based on the ML outcomes, the vaccine efficacy was calculated against the unvaccinated group. Protection efficacy of 55% (95%CI 43%–64%) was obtained with two doses of vaccine. This was per the available literature. From recently published work of Covaxin Phase 3 trial, 63·6% (95%CI 29·0%–82·4%) protection efficacy was against asymptomatic infection while 65·2% (95%CI 33·1%–83·0%) protection was observed against the Delta variant (Ella et al., 2021).

## Discussion

The systemic immune response elicited by the whole virion vaccine may be challenging to distinguish vaccine immunogenicity from the pathogen infection response of the host. Based on the analysis of Anti-NC and Anti-S antibody response and the number of days since the last vaccination from the serosurvey-based data of Covaxin administered individuals, we identified two subgroups of people agnostic of their self-reported infection status. Though one subgroup was highly enriched in self-reported positive people, the self-reported negative individuals were distributed throughout the two subgroups. We hypothesized that the self-reported negative individuals who are in the same group as that of self-reported positive people may have been infected earlier, irrespective of self-reporting to be negative. This could be because of multiple reasons e.g. a person might have got infected after the test, false-negative report of the RT-PCR, etc.

To collect evidence of infection for negative RT-PCR people, we obtained the seropositivity status in the previous phases of the serosurvey cohort along with the self-reported infection status followed by the supervised and the unsupervised machine learning approaches. The consensus was built for 90.8% of them (965 out of 1063) individuals for infection and non-infection status. To further validate the infection status proposed by our pipeline, the sVNT assays performed on samples from Phase 3 individuals resulted in the identification of Delta infected individuals who were self-reported as uninfected. The Delta Infected samples were highly correspondent with the ML predicted Infected people thereby reinforcing our hypothesis. To make use of this method, we provide a machine learning method based on 0.99 AUROC on the updated infection status to predict the new Covaxin administered participants for a prior infection status.

Though this study fills the gap in the field, it comes with certain limitations. First, the self-reported status is questionnaire-based which might have a certain level of inconsistency. Second, not infected individuals might get infected at any time after the questionnaire form filling, which means that longitudinal data might have been better at capturing the real infection rate. Third, the samples come from different employees of institutes and their relatives, which might not be the real representation of the overall country’s population, especially in rural areas.

The geographical locations covered by our study include CSIR labs located throughout the country (Naushin et al., 2021) which complements the predominantly rural locale by the ICMR study. Further, a recent ICMR paper of pilot study with 114 individuals shows that people with infection and 1 dose of Covaxin are equally responsive to the ones with infection naïve 2 doses (Kumar et al., 2021). This study was able to address an important gap in available literature for calculating vaccine effectiveness with whole virion vaccine from serology-based surveys. Thus, our work fills the gap where we developed and validated a method to identify asymptomatic COVID-19 infected individuals and thus were able to calculate vaccine efficacy in one of the largest cohorts of India. Finally, we were able to provide the protection efficacy (PE) of 55% (95%CI 43%–64%) for fully vaccinated subjects. This was per the available literature where 63·6% (95%CI 29·0%–82·4%) protection efficacy was observed against asymptomatic infection after two doses of vaccine (Ella et al., 2021), which were phase 3 trial results. The vaccine efficacy provided by our model was very similar to the recently published real-world efficacy of 47% (95%CI 29%–61%) after two doses in previously uninfected individuals who had symptomatic presentation (Desai et al., 2021). This proved the importance of our work and how the ML-based approach could be utilized to study real-world effectiveness even in asymptomatic population-based on serological methods.

## Conclusion

Through induction of Anti-NC, whole virion vaccines, such as BBV152, pose a difficulty in identifying an infection status for serological surveys. As RT-PCR may miss significant asymptomatic infection, we developed an ML-based approach, to predict prior infection status in partially and fully vaccinated Covaxin recipients with significant accuracy. Using the ML-based approach, we could observe a PE of 55% (95%CI 43%–64%) in fully vaccinated subjects which agreed with available literature. This work fills an important gap in identifying infection status in whole virion vaccine recipients through serological data in conjunction with an ML-based approach.

## Data Availability

All data produced in the present study are available upon reasonable request to the authors

## Acknowledgment

SSG would like to acknowledge CSIR grant MLP 2007 for this work. Salwa Naushin and Rajat Ujjainiya would like to thank CSIR for their fellowships. Prateek Singh would like to thank DBT-BINC for their fellowship. Satyartha Prakash would like to acknowledge CSIR grant MLP-1811(CSIR-IGIB) for the fellowship. AA, VS, and Nitin Bhatheja would like to acknowledge DST grant GAP-0192 (CSIR-IGIB) in partial support of this work. Ajay Pratap Singh would like to address CSIR grant MLP-2002 (CSIR-IGIB) for this work. We would also like to thank the Directors of all the CSIR Institutes for facilitating the study. The authors also thank Pushpesh Ranjan, Neeraj Kumar, Shalu Yadav, Ayushi Singhal, and Md. Abubakar Sadique from AMPRI. V Santosh Kumar from CCMB. Dr. Shalini Gupta, Shraddha, and Nandpal Yadav from CDRI. PK Khanna, Vipul Sharma, Hansraj Choudhary, Nalini Pareek, Maninder Kaur, Vijay Chatterjee, RK Singh, Prem Kumar, Narendra Meena, Ramesh Baura, Virender Singh, Mahendra Singh, SK Masiul Islam, Anuroop Bajpai, and Khusbhu from CEERI. Krishna Kommatwar, Meena Kumari, Monisha Vijayrengan, RamyaR, Dr. Avilash S, Rani C, and Naveen Shashidhar Kumbar from CFTRI. DC Sharma, Dr. Neelam J Gupta, AK Jain and Sudhansu Bhagat from CRRI. Deepesh Khandwal, Shrikant Khandare and Nirmala Kumari Gupta from CSMCRI. Ajeet Singh and Dr. RK Sinha from HRDC. Dr. Sanghamitra Pati from ICMR-RMRC. Pankaj Pandey and Rajesh from IGIB. Jaideep Mehta, Dinesh Kumar, Dr. Dhan Prakash, Vineet Kumar, Rajendra Dass, Dr. Raj Kumar, Sumit Mittal, Deepak Bhat, Paramjit Kashyap, Anjali Koundal, and Dr. Sanjeev Khosla from IMTECH. Sandeep Negi from IITR. G Bhatt, Shashikala U, Shashidhar K N, Dr. Amarnarayan D from NAL. Ravichandran C, RP Thagavelu and Sunil Babu M G from 4PI. Dr. Y Madhavi from NIScPR. Dr. Sumit Dahiya from NPL. Dr. Robin Joshi, Dr. Anish Kaachra, Dr. Pankaj Kulurkar from IHBT. Masood Ahmed, Nitika Kapoor, Chahat Chopra, Rekha Chouhan, Loveleenakaur Anand from IIIM. Megha Sailwal, Ayan Banerjee, Neha Bansal, Abhilek Nautiyal from IIP. Naveen Kumar from KR Diagnostic. Dr. Meganathan P. R. and Dr. Shaik Basha from NEERI-Hyderabad. Thaneswar Bora from NEIST. Dr. Shuchismita Benzwal and Dr. Chaitanya Dinesh from NGRI. Shana S Nair and Valan Rebinro from NIIST. Team NML. Dr. S Maheswaran, Dr. N. Anandavalli and P Vasudevan from SERC. Rashmi Arya from URDIP.

## References

Bai, Y., Yao, L., Wei, T., Tian, F., Jin, D.Y., Chen, L., and Wang, M. (2020). Presumed Asymptomatic Carrier Transmission of COVID-19. JAMA 323, 1406–1407.

Desai, D., Khan, A.R., Soneja, M., Mittal, A., Naik, S., Kodan, P., Mandal, A., Maher, G.T., Kumar, R., Agarwal, A., et al. (2021). Effectiveness of an inactivated virus-based SARS-CoV-2 vaccine, BBV152, in India: a test-negative, case-control study. Lancet Infect Dis.

Dhar, M.S., Marwal, R., Vs, R., Ponnusamy, K., Jolly, B., Bhoyar, R.C., Sardana, V., Naushin, S., Rophina, M., Mellan, T.A., et al. (2021). Genomic characterization and epidemiology of an emerging SARS-CoV-2 variant in Delhi, India. Science, eabj9932.

Ella, R., Reddy, S., Blackwelder, W., Potdar, V., Yadav, P., Sarangi, V., Aileni, V.K., Kanungo, S., Rai, S., Reddy, P., et al. (2021). Efficacy, safety, and lot-to-lot immunogenicity of an inactivated SARS-CoV-2 vaccine (BBV152): interim results of a randomised, double-blind, controlled, phase 3 trial. Lancet.

Estiri, H., Strasser, Z.H., Klann, J.G., Naseri, P., Wagholikar, K.B., and Murphy, S.N. (2021). Predicting COVID-19 mortality with electronic medical records. NPJ Digit Med 4, 15.

Gupta, R.K., Marks, M., Samuels, T.H.A., Luintel, A., Rampling, T., Chowdhury, H., Quartagno, M., Nair, A., Lipman, M., Abubakar, I., et al. (2020). Systematic evaluation and external validation of 22 prognostic models among hospitalised adults with COVID-19: an observational cohort study. Eur Respir J 56.

Hodgson, S.H., Mansatta, K., Mallett, G., Harris, V., Emary, K.R.W., and Pollard, A.J. (2021). What defines an efficacious COVID-19 vaccine? A review of the challenges assessing the clinical efficacy of vaccines against SARS-CoV-2. Lancet Infect Dis 21, e26–e35.

Kang, Y.M., Choe, K.W., Lee, K.D., Kim, K.N., Kim, M.J., and Lim, J. (2021). SARS-CoV-2 Antibody Response to the BNT162b2 mRNA Vaccine in Persons with Past Natural Infection. J Korean Med Sci 36, e250.

Kumar, N.P., Padmapriyadarsini, C., Uma Devi, K.R., Banurekha, V.V., Nancy, A., Girish Kumar, C.P., Murhekar, M.V., Gupta, N., Panda, S., Babu, S., et al. (2021). Antibody responses to the BBV152 vaccine in individuals previously infected with SARS-CoV-2: A pilot study. Indian J Med Res 153, 671–676.

Lee, C.Y., Lin, R.T.P., Renia, L., and Ng, L.F.P. (2020). Serological Approaches for COVID-19: Epidemiologic Perspective on Surveillance and Control. Front Immunol 11, 879.

Mallett, S., Allen, A.J., Graziadio, S., Taylor, S.A., Sakai, N.S., Green, K., Suklan, J., Hyde, C., Shinkins, B., Zhelev, Z., et al. (2020). At what times during infection is SARS-CoV-2 detectable and no longer detectable using RT-PCR-based tests? A systematic review of individual participant data. BMC Med 18, 346.

Marbac, M., and Sedki, M. (2019). VarSelLCM: an R/C++ package for variable selection in model-based clustering of mixed-data with missing values. Bioinformatics 35, 1255–1257.

Naushin, S., Sardana, V., Ujjainiya, R., Bhatheja, N., Kutum, R., Bhaskar, A.K., Pradhan, S., Prakash, S., Khan, R., Rawat, B.S., et al. (2021). Insights from a Pan India Sero-Epidemiological survey (Phenome-India Cohort) for SARS-CoV2. Elife 10.

Nishiura, H., Kobayashi, T., Miyama, T., Suzuki, A., Jung, S.M., Hayashi, K., Kinoshita, R., Yang, Y., Yuan, B., Akhmetzhanov, A.R., et al. (2020). Estimation of the asymptomatic ratio of novel coronavirus infections (COVID-19). Int J Infect Dis 94, 154–155.

Oran, D.P., and Topol, E.J. (2020). Prevalence of Asymptomatic SARS-CoV-2 Infection : A Narrative Review. Ann Intern Med 173, 362–367.

Pedregosa, F., Varoquaux, G., Gramfort, A., Michel, V., Thirion, B., Grisel, O., Blondel, M., Prettenhofer, P., Weiss, R., and Dubourg, V. (2011). Scikit-learn: Machine learning in Python. the Journal of machine Learning research 12, 2825–2830.

Pelleau, S., Woudenberg, T., Rosado, J., Donnadieu, F., Garcia, L., Obadia, T., Gardais, S., Elgharbawy, Y., Velay, A., and Gonzalez, M. (2021). Serological reconstruction of COVID-19 epidemics through analysis of antibody kinetics to SARS-CoV-2 proteins. medRxiv.

Sapkal, G., Yadav, P.D., Ella, R., Abraham, P., Patil, D.Y., Gupta, N., Panda, S., Mohan, V.K., and Bhargava, B. (2021a). Neutralization of VUI B.1.1.28 P2 variant with sera of COVID-19 recovered cases and recipients of Covaxin an inactivated COVID-19 vaccine. J Travel Med 28.

Sapkal, G.N., Yadav, P.D., Ella, R., Deshpande, G.R., Sahay, R.R., Gupta, N., Vadrevu, K.M., Abraham, P., Panda, S., and Bhargava, B. (2021b). Inactivated COVID-19 vaccine BBV152/COVAXIN effectively neutralizes recently emerged B.1.1.7 variant of SARS-CoV-2. J Travel Med 28.

Singanayagam, A., Hakki, S., Dunning, J., Madon, K.J., Crone, M.A., Koycheva, A., Derqui-Fernandez, N., Barnett, J.L., Whitfield, M.G., Varro, R., et al. (2021). Community transmission and viral load kinetics of the SARS-CoV-2 delta (B.1.617.2) variant in vaccinated and unvaccinated individuals in the UK: a prospective, longitudinal, cohort study. Lancet Infect Dis.

Szepannek, G. (2018). clustMixType: User-Friendly Clustering of Mixed-Type Data in R. R J 10, 200.

Yadav, P.D., Sapkal, G.N., Abraham, P., Ella, R., Deshpande, G., Patil, D.Y., Nyayanit, D.A., Gupta, N., Sahay, R.R., Shete, A.M., et al. (2021a). Neutralization of variant under investigation B.1.617 with sera of BBV152 vaccinees. Clin Infect Dis.

Yadav, P.D., Sapkal, G.N., Ella, R., Sahay, R.R., Nyayanit, D.A., Patil, D.Y., Deshpande, G., Shete, A.M., Gupta, N., Mohan, V.K., et al. (2021b). Neutralization of Beta and Delta variant with sera of COVID-19 recovered cases and vaccinees of inactivated COVID-19 vaccine BBV152/Covaxin. J Travel Med 28.

Zoabi, Y., Deri-Rozov, S., and Shomron, N. (2021). Machine learning-based prediction of COVID-19 diagnosis based on symptoms. NPJ Digit Med 4, 3.

