## Supplementary figures and tables for "A machine learning-based approach to determine infection status in recipients of BBV152 whole virion inactivated SARS-CoV-2 vaccine for serological surveys"

#### Supplementary Tables

**Table S1:** Distribution of Covaxin-administered individuals across 43 Institutes/Centers.

| Lab/Center | Sample Count | Lab/Center | Sample Count | Lab/Center | Sample count | Lab/Center | Sample count |
|---|---|---|---|---|---|---|---|
| CSIR - IICT | 348 | CSIR - NPL | 34 | CSIR - NCL | 18 | CSIR - TKDL | 12 |
| CSIR - IMMT | 270 | CSIR - CRRI | 30 | CSIR - IITR | 17 | CSIR - AMPRI | 11 |
| CSIR - NAL | 211 | CSIR - HQ | 29 | CSIR – Apartments | 16 | CSIR - CBRI | 8 |
| CSIR - CFTRI | 110 | CSIR – IIIM, Jammu | 28 | CSIR - CIMAP | 16 | CSIR - CGCRI | 8 |
| CSIR - CLRI | 95 | CSIR - NML | 28 | CSIR - IICB | 15 | CSIR - HRDC | 8 |
| CSIR - IGIB | 69 | CSIR - IMTECH | 27 | CSIR - NBRI | 15 | CSIR - IHBT | 4 |
| CSIR - SERC | 52 | CSIR - NIScPR | 22 | CSIR - PUSA | 14 | CSIR - CMERI | 3 |
| CSIR - CDRI | 49 | CSIR - IIP | 20 | CSIR - CSIO | 13 | CSIR - CIMFR | 2 |
| CSIR - CEERI | 45 | CSIR - NGRI | 19 | CSIR - NIO | 13 | CSIR - URDIP | 2 |
| CSIR - NEERI | 42 | CSIR - NIIST | 19 | CSIR - NPL Colony | 13 | CSIR - IIIM, Srinagar | 1 |
| CSIR - CCMB | 37 | CSIR - CSMCRI | 18 | CSIR - CECRI | 12 | | |

**Table S2:** Baseline demographic characteristics of all individuals, also stratified as 1 Dose and 2 Doses.

| Group | | Overall | 1 Dose | 2 Doses | P-value |
|---|---|---|---|---|---|
| | | 1823 | 772 (42.3%) | 1051 (57.6%) | |
| Gender N (%) | Female | 609 (33.4) | 242 (31.3) | 367 (34.9) | 0.12 |
| | Male | 1214 (66.6) | 530 (68.7) | 684 (65.1) | |
| Age (Median (IQR)) | | 42 (31-50) | 39 (30-46) | 45 (33-53) | <0.05 |

### Supplementary Figures

(A)

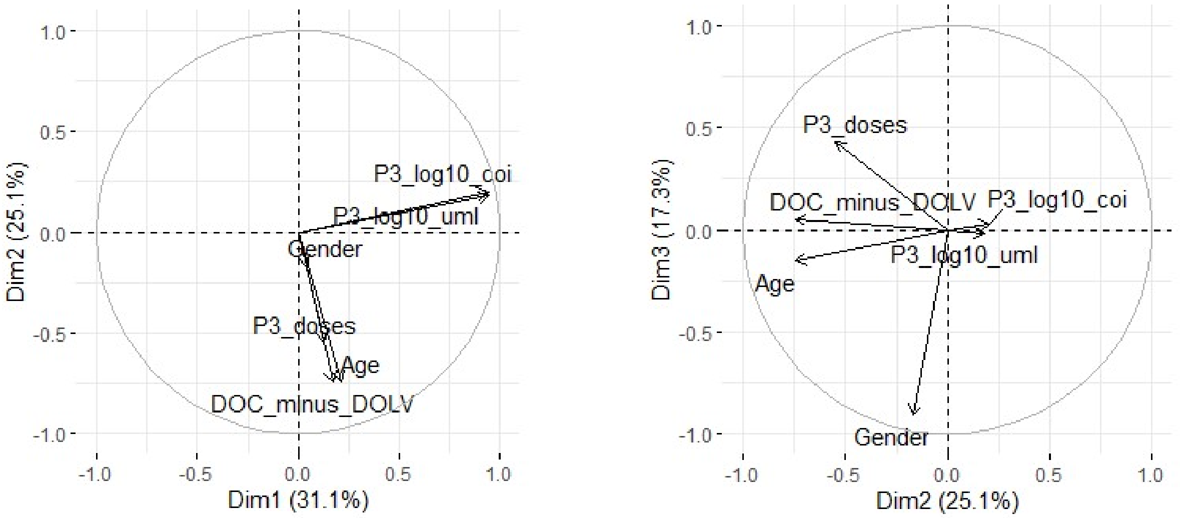


(B)

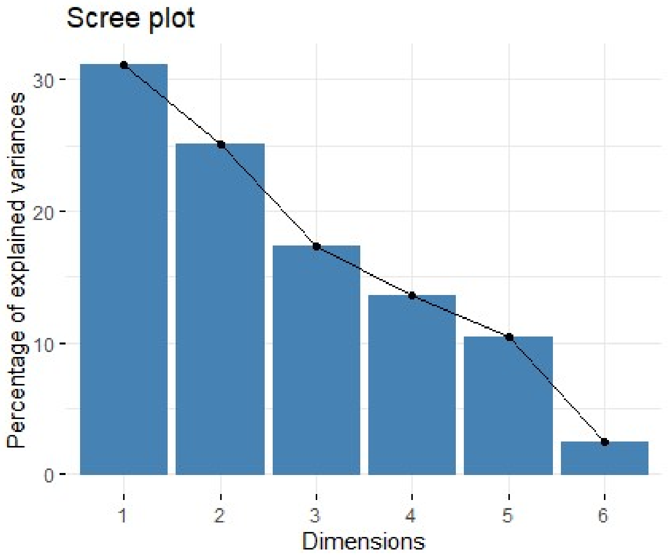


**Figure S1:** A) *Variable correlation plot of six parameters used to perform PCA analysis. COI and U/mL are correlated while the number of doses and days since the last vaccination are grouped. Gender has a contribution to the variance in the PC3 dimension. B) Scree plot showcasing percentage variance explained by each Principal Component. The first three principal components together explain 73.5% of the variance in the data.*

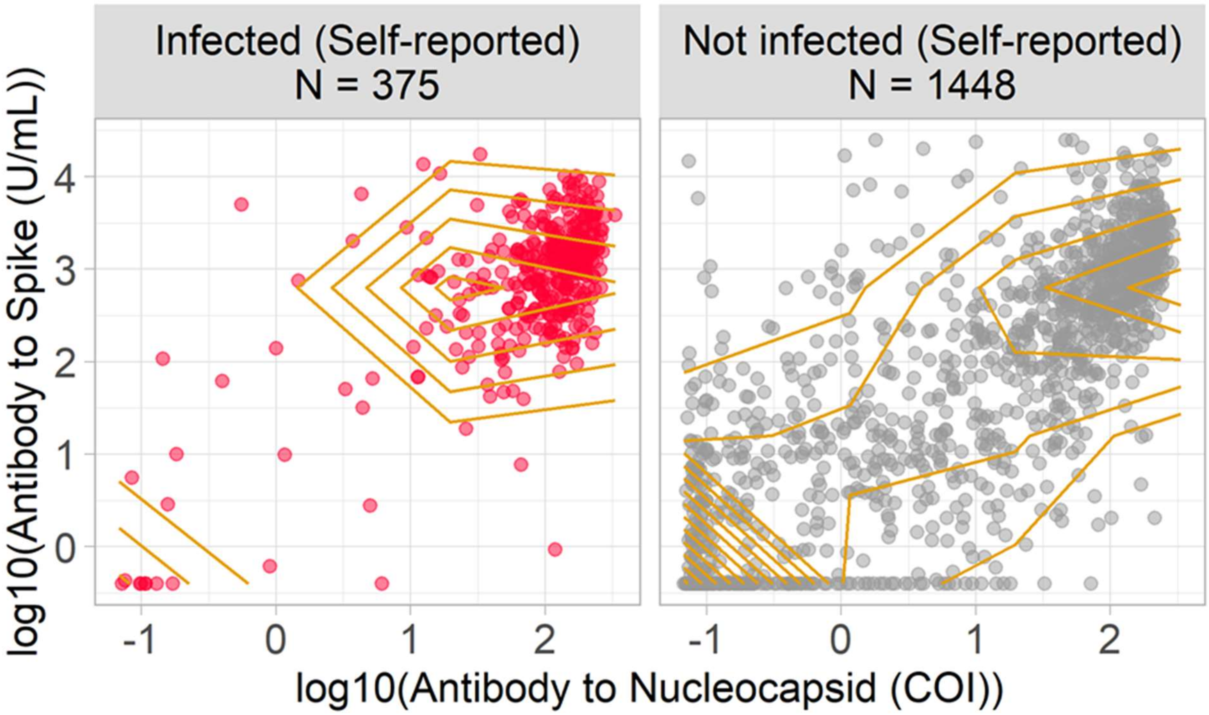


**Figure S2:** Sample distribution stratified via self-reported COVID-19 infection status. Density-based contours indicate the presence of two subgroups amongst the self-reported not infected individuals.

(A)

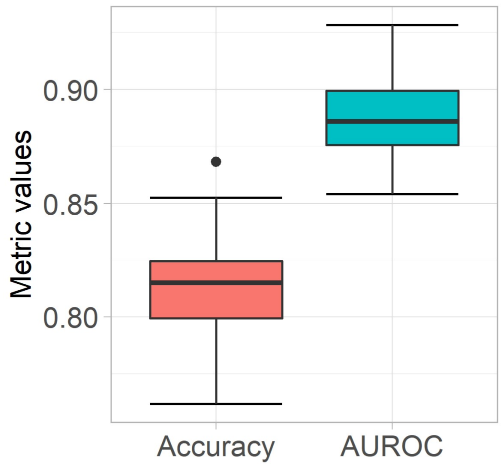


(B)

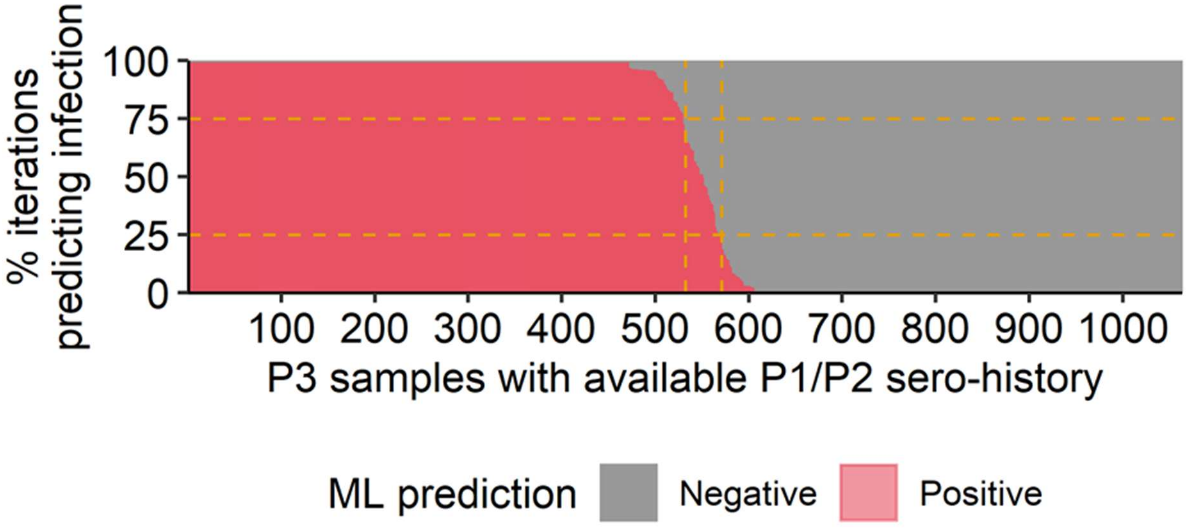


**Figure S3: A)** Average ML-based (SVM) model performance evaluation metrics results for 100 iterations (N=1063). **B)** Cutoff selection after 100 iterations. <25% = Not infected, >75% = infected, 25%-75% = Indeterminate.

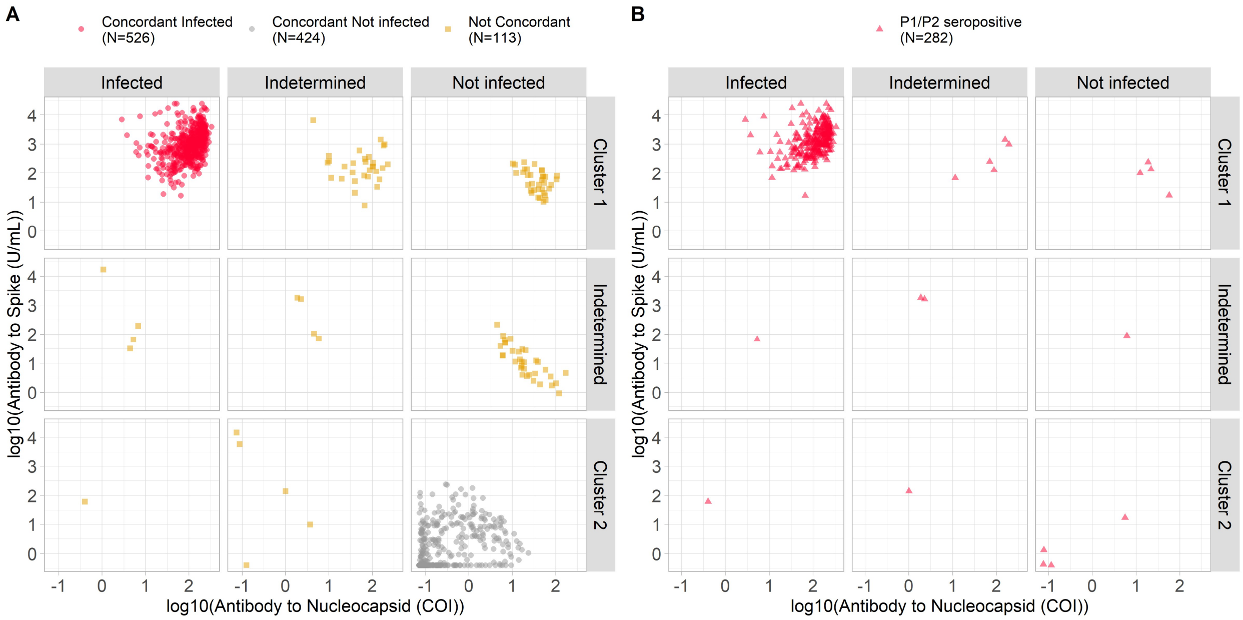


**Figure S4:** Concordance between SVM-based model and Consensus Clustering approaches on the Covaxin administered people (N=1063). For the ensemble model, we used concordant samples (red (N=526) and grey color (N=424) in Panel A) along with samples that were seropositive in P1/P2 (i.e. red triangles in Panel B, N=15)

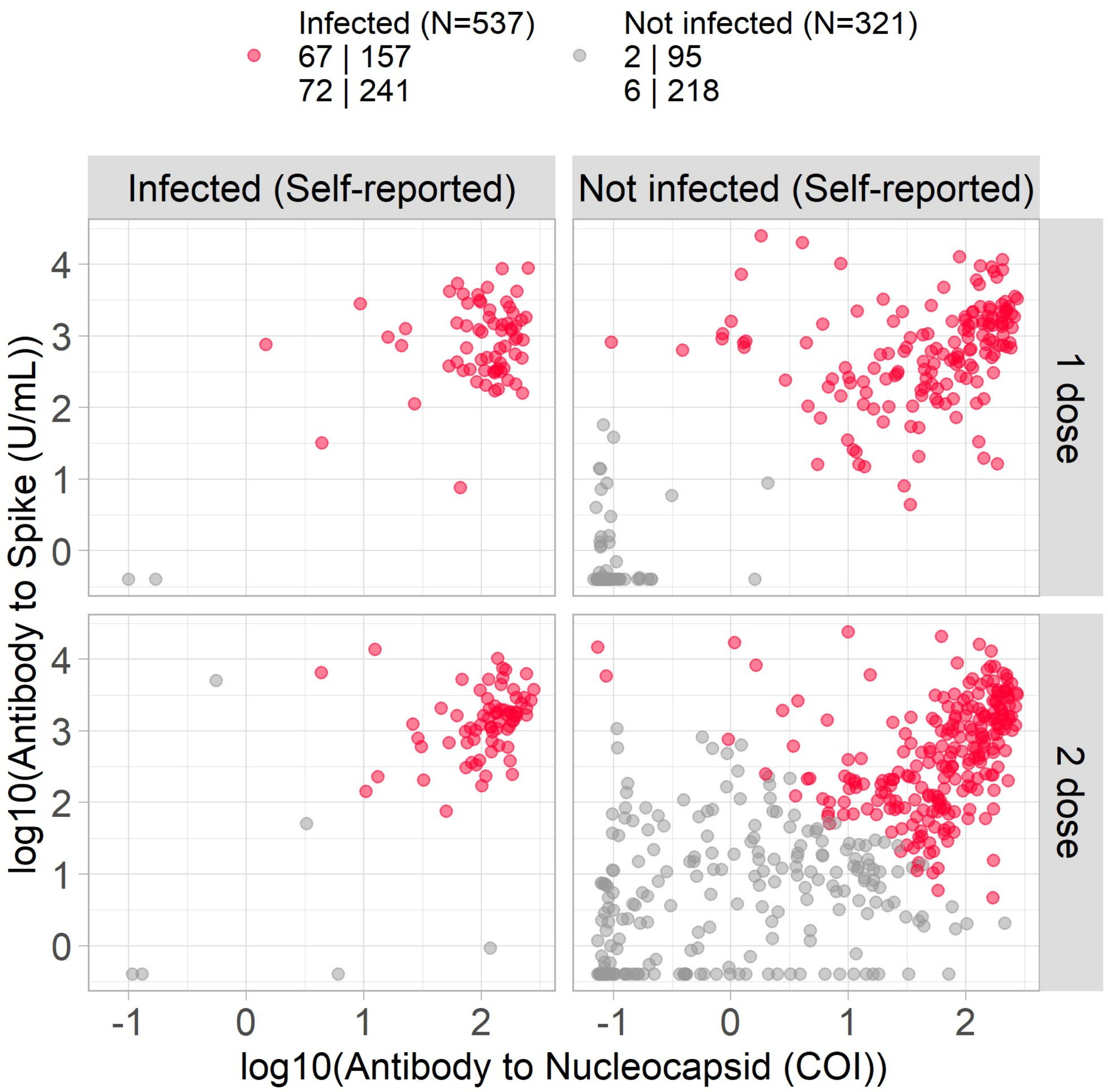


**Figure S5:** Final Ensemble ML model-based prediction of the infection status of leftover samples (N=858) not used in the model building, further stratified via self-reported infection status and the number of vaccine doses.

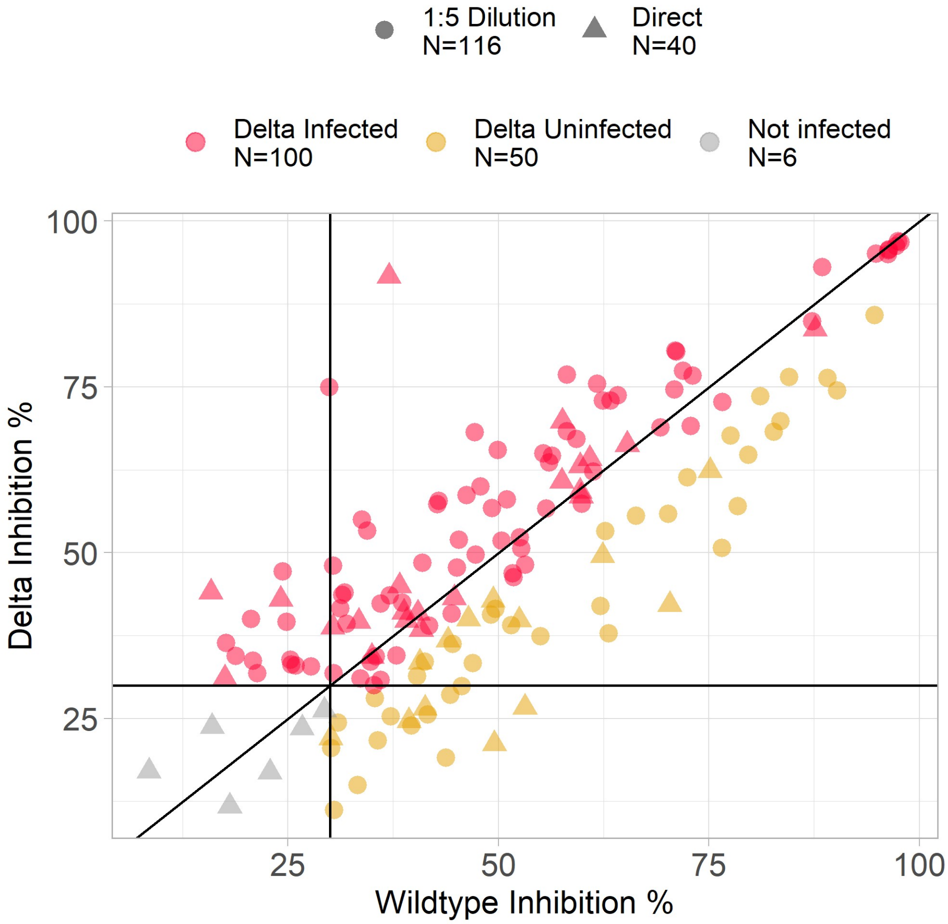


**Figure S6:** A surrogate virus neutralization assay (sVNT) of P3 individuals who reported to be COVID-19 negative and predicted to be Infected by Ensemble model (N=156). 64.1% of samples predicted to be Infected by Ensemble were found to be Delta Infected by sVNT. Delta Infected was labelled when Delta Inhibition % > WT Inhibition % with a margin based on standard error. Delta Not infected were labelled when samples processed without dilution had less than 30% inhibition. All other data points were labeled Delta Uninfected.
